# Cross-Source Progression Assessment in Advanced Pancreatic Cancer: An Opportunity-Adjusted Six-State Analysis

**DOI:** 10.64898/2026.09.13.26362944

**Authors:** Hamzah Atef Jaareh, Farah Habash, Salaheddin Mustafa, Eyas Hasan Shaker Saleh

**Author notes:** Corresponding author **Corresponding author** Hamzah Atef Jaareh, MD, Shmaisani Hospital, Amman, Jordan. Competing interests The authors declare no competing interests. **Author contributions** Conception and design: All authors. Collection and assembly of data: All authors. Data analysis and interpretation: All authors. Manuscript writing: All authors. Final approval of manuscript: All authors.

## Abstract

**PURPOSE:** Radiology and treating-oncologist documentation are recorded on different schedules in routine oncology care. We characterized the opposite clinical source at its first evaluable opportunity after radiology- or medical-oncology-documented progression in advanced pancreatic cancer using an exhaustive six-state framework.

**METHODS:** We performed a retrospective secondary analysis of AACR Project GENIE BPC PANC v1.0-public. Patients entered the treatment-index cohort at the first systemic regimen initiated on or after advanced-disease onset. Process T0 was the first genuine radiology- or medical-oncology-documented progression after cohort entry and not after subsequent same-cancer treatment or death. The opposite source’s first evaluable opportunity was classified as same-day concordance, confirmation, explicit nonprogression, mixed, indeterminate, or no evaluable opportunity. The secondary estimand was explicit nonprogression among first opportunities classifiable specifically as progression or explicit nonprogression.

**RESULTS:** Of 1,109 patients, 882 entered the treatment-index cohort and 645 qualified for the primary estimand. States were same-day concordance in 25 (3.88%), confirmation in 219 (33.95%), explicit nonprogression in 108 (16.74%), mixed in 19 (2.95%), indeterminate in 166 (25.74%), and no evaluable opportunity in 108 (16.74%). Among 327 progression/nonprogression-classifiable first opportunities, 108 (33.03%; 95% CI, 28.15%-38.30%) documented explicit nonprogression. Opposite-source opportunity occurred after 88.47% of imaging-first events at a median of 5 days versus 62.94% of medical-oncology-first events at a median of 38 days. A known-regimen-completion sensitivity yielded 84/144 (58.33%).

**CONCLUSION:** Cross-source progression assessment frequently did not yield immediate confirmation and often remained indeterminate or unevaluable. An opportunity-adjusted six-state framework makes observation-process and clinical-boundary dependencies explicit rather than compressing them into a binary discordance estimate.

## Introduction

During systemic therapy for advanced pancreatic cancer, disease status is reassessed longitudinally using clinical evaluation and serial imaging. [16-18] CA19-9 may add context, but it cannot substitute for imaging or serve as a stand-alone truth standard because of important biological and clinical limitations. [16,20]

In routinely collected oncology data, progression is not continuously observed. Radiology reports and clinician documentation can both support real-world progression endpoints, but neither reproduces a protocolized RECIST assessment schedule, and both are exposed to intermittent observation and differential assessment frequency. [5-6,8-11] Consequently, the recorded timing and apparent source of progression can depend on when each source is observed.

Radiology-anchored and clinician-anchored real-world progression have already been compared, and AACR Project GENIE BPC studies have defined imaging-based PFS (PFS-I), medical-oncology-based PFS (PFSM), and combined variants. [5,4,3] The remaining methodological problem is not whether the sources can differ, but how to describe their relationship without conditioning the analysis on patients whose opposite-source assessment happens to be timely and classifiable.

AACR Project GENIE BPC PANC v1.0-public separately curates longitudinal imaging and medical-oncology status, allowing this cross-source process to be reconstructed directly. [1-2] We anchored patients at the first progression documented by either source after initiation of the first advanced systemic regimen and before subsequent same-cancer treatment or death, then classified the opposite source at its first evaluable opportunity.

A prespecified literature search through September 9, 2026 identified pancreatic cancer studies of real-world progression, treatment decisions after radiographic progression, serial CT prediction, and genomic outcomes in the same dataset family, but none reproduced the complete source-first, first-opposite-source-opportunity six-state framework used here. [12-15] We therefore estimated the full state distribution, quantified source-specific observation opportunity, and tested robustness to alternative endpoint and clinical-boundary definitions. The contribution is the integrated measurement-process framework, not the underlying concept of radiology-versus-clinician progression or PFS-I/PFS-M. [5,4]

## Methods

### Study design and data source

We performed a retrospective secondary analysis of the AACR Project GENIE Biopharma Collaborative pancreatic cancer cohort (BPC PANC v1.0-public), a multi-institutional longitudinal clinicogenomic resource with separately curated imaging and medical-oncology assessments. The analytic release includes 1,109 patients from four institutions. AACR Project GENIE is an international data-sharing consortium that releases deidentified clinico-genomic data for research. [27] Variable definitions and longitudinal source semantics were taken from the official PANC analytic documentation, and reporting was planned in accordance with STROBE and RECORD principles for observational studies using routinely collected health data. [1-2,25-26]

Human investigations. This retrospective secondary analysis used only deidentified AACR Project GENIE BPC PANC v1.0-public data. The investigators had no interaction or intervention with participants and no access to direct identifiers. AACR Project GENIE contributing institutions protect patient privacy under applicable local consent and institutional-review frameworks, including consent or institutional review board waiver as appropriate. [27] Because the present study was limited to secondary analysis of deidentified data released for research use, additional institutional review board review and individual informed consent were not required for this analysis.

### Treatment-index cohort

We restricted the source population to the index pancreatic cancer and defined advanced disease using the official derived PFS cohort categories Stage IV or Stage I-III with Distant Mets. For each patient, regimen rows were linked to the same index cancer, and cohort entry was the first cancer-directed systemic regimen whose start occurred on or after advanced-disease onset. All dates were harmonized on the DOB-relative day scale. Regimen start reconstructed from cancer-diagnosis-relative intervals was cross-checked against absolute drug-start intervals before analysis. [1-2]

### Source-specific progression reconstruction

We reconstructed source-specific progression from the raw curated status fields rather than from composite PFS event flags. Radiology used image_overall and medical oncology used md_ca_status; the PANC data guide documents both variables and specifies that PFS-I and PFS-M composite event flags can include progression or death. Composite PFS flags were therefore not treated as pure source-specific progression indicators. Progressing/Worsening/Enlarging mapped to PROGRESSION, Mixed to MIXED, Stable/No change or Improving/Responding to NONPROGRESSIVE, and Not stated/Indeterminate or otherwise nonclassifiable assessments to INDETERMINATE. These were analyzed as curated source states, not as a centralized RECIST 1.1 rereview. [2,4,21]

### Opportunity, six-state outcome, and clinical boundaries

Assessments from the same source on the same calendar day were collapsed before cross-source comparison. For imaging, the prespecified hierarchy was PROGRESSION > MIXED > NONPROGRESSIVE > INDETERMINATE; stricter treatment of same-day conflicts was examined in sensitivity analysis. T0 was the earliest day on or after cohort entry on which either source documented day-level PROGRESSION, provided that day was not after the first subsequent same-cancer regimen start or death. The index source was imaging first, medical oncology first, or same-day dual-source progression. The first cross-source opportunity was the earliest assessment by the opposite source on or after T0 and before the clinical boundary or administrative close. If an opposite-source assessment and a clinical boundary occurred on the same day and ordering could not be resolved, that opportunity was classified as indeterminate. Each qualifying patient contributed exactly one state: SAME_DAY_CONCORDANCE, CONFIRMED_AT_FIRST_OPPORTUNITY, EXPLICIT_NONPROGRESSION, MIXED, INDETERMINATE, or NO_EVALUABLE_CROSS_SOURCE_OPPORTUNITY. Documented completion of all drugs in a regimen (dx_reg_end_all_int) was used only in a prespecified sensitivity analysis. [2]

### Primary and secondary estimands

The primary estimand was the six-component state distribution among all patients with qualifying source-specific progression, reported as counts, percentages of the full primary denominator, and 95% Wilson score confidence intervals. The secondary conditional estimand was the proportion with EXPLICIT_NONPROGRESSION among first opportunities classifiable specifically as progression or explicit nonprogression. Mixed, indeterminate, same-day-concordant, and no-opportunity states remained part of the primary distribution rather than being excluded by conditioning. [23]

### Observation-process timing and competing risks

We summarized T0-to-first-opposite-source opportunity and T0-to-opposite-source progression confirmation with medians and interquartile ranges. Cumulative incidence of opposite-source confirmation at 14, 30, 60, and 90 days was estimated with the Aalen-Johansen method, treating subsequent same-cancer treatment or death as competing events and administrative close as censoring. [22]

### Clinical-context analyses

Subsequent treatment was reconstructed from the actual next same-index-cancer regimen row rather than a composite time-to-next-treatment indicator. CA19-9 was analyzed only as contextual longitudinal information, not as a biological adjudicator of either source. After unit quality control, the prespecified primary contextual windows selected the closest valid CA19-9 value in days -60 to -1 and the closest valid value in days 0 to +60 relative to T0; paired change was summarized continuously using raw change and log1p(post) - log1p(pre), without a response threshold. Living patients in descriptive post-T0 overall-survival analyses were censored at the official composite last-known-alive variable dob_lastalive_int, and survival was estimated by Kaplan-Meier methods. [2,16,19-20,24]

### Supportive adjusted analysis and sensitivity analyses

A prespecified logistic model was fit among first opportunities classifiable as confirmation or explicit nonprogression, with explicit nonprogression as the modeled event. The fixed covariates were age, sex, institution, advanced presentation, first advanced-regimen class, index source, and log-transformed pre-T0 imaging and medical-oncology assessment counts; model coefficients were treated as secondary, supportive, and noncausal. Sensitivity analyses examined a single-primary-cancer restriction, stricter same-day conflict handling, counting MIXED as confirmation, permissive boundary-day ordering, stricter cross-source evaluability, a known-regimen-completion boundary using documented dx_reg_end_all_int, prespecified clinical and observation-density strata, confirmation windows, alternative CA19-9 windows, and a boundary-aware CA19-9 analysis. Because progression ascertainment is sensitive to assessment timing and frequency, source-observation opportunity was analyzed as part of the measurement process rather than treated as ignorable missingness. [2,8-11]

Artificial intelligence use in the research workflow. OpenAI GPT-5.6 Sol (OpenAI; accessed September 9, 2026) was used as a research-assistance tool for analytic workflow planning, code generation and execution orchestration, structured quality-control and reconstruction checks, literature/source verification, and preparation of tables, figures, and manuscript text. Statistical results reported here were generated or regenerated by deterministic code and subjected to prespecified validation and cross-checking. The AI system was not treated as an author or autonomous clinical adjudicator. The human authors reviewed and approved all analytic definitions, results, interpretations, citations, and final text and retain full responsibility for the work.

## Results

### Cohort and primary six-state distribution

Of 1,109 patients in the PANC release, 1,021 had advanced index pancreatic cancer, 958 had an index-cancer regimen row, and 882 initiated an eligible cancer-directed regimen on or after advanced-disease onset (Figure 1). [1] Among these 882 patients, 645 had at least one qualifying genuine source-specific progression after cohort entry and not after the next same-cancer treatment/death boundary. Median age at cohort entry in the 645-patient progression-process cohort was 65.3 years (IQR, 57.9-71.5); 301 (46.7%) were female, 323 (50.1%) presented with de novo stage IV disease, and 516 (80.0%) had one recorded primary cancer (Table 1). First genuine progression was imaging-first in 477 patients (74.0%), medical-oncology-first in 143 (22.2%), and same-day dual-source in 25 (3.9%).

**Table 1.**

| Section | Characteristic | Value | Denominator |
| --- | --- | --- | --- |
| Demographics | Age at cohort entry, median (IQR), years | 65.3 (57.9–71.5) | 645 |
| Demographics | Sex: Female | 301 (46.7%) | 645 |
| Demographics | Sex: Male | 344 (53.3%) | 645 |
| Care setting | Institution: DFCI | 245 (38.0%) | 645 |
| Care setting | Institution: MSK | 309 (47.9%) | 645 |
| Care setting | Institution: UHN | 46 (7.1%) | 645 |
| Care setting | Institution: VICC | 45 (7.0%) | 645 |
| Disease presentation | De novo Stage IV | 323 (50.1%) | 645 |
| Disease presentation | Stage I-III then distant mets | 322 (49.9%) | 645 |
| First advanced regimen | FOLFIRINOX/mFOLFIRINOX-containing | 277 (42.9%) | 645 |
| First advanced regimen | Gemcitabine+nab-paclitaxel | 175 (27.1%) | 645 |
| First advanced regimen | Other gemcitabine-based | 111 (17.2%) | 645 |
| First advanced regimen | Fluoropyrimidine/oxaliplatin-based | 28 (4.3%) | 645 |
| First advanced regimen | Other | 54 (8.4%) | 645 |
| Cancer history | Recorded primary cancers: 1 cancer | 516 (80.0%) | 645 |
| Cancer history | Recorded primary cancers: >1 cancer | 129 (20.0%) | 645 |
| Cancer history | Index-cancer systemic treatment before advanced onset: No | 428 (66.4%) | 645 |
| Cancer history | Index-cancer systemic treatment before advanced onset: Yes | 217 (33.6%) | 645 |
| Observation process | First genuine progression source: Imaging first | 477 (74.0%) | 645 |
| Observation process | First genuine progression source: Medical oncology first | 143 (22.2%) | 645 |
| Observation process | First genuine progression source: Same-day dual-source | 25 (3.9%) | 645 |
| Observation process | Pre-T0 imaging assessments, median (IQR) | 1 (0–3) | 645 |
| Observation process | Pre-T0 med-onc assessments, median (IQR) | 2 (1–5) | 645 |
| Observation process | Observed opposite-source opportunity | 537 (83.3%) | 645 |
| Biomarker context | Paired CA19-9 evaluable in prespecified ±60-day windows | 328 (50.9%) | 645 |

**Figure 1.**
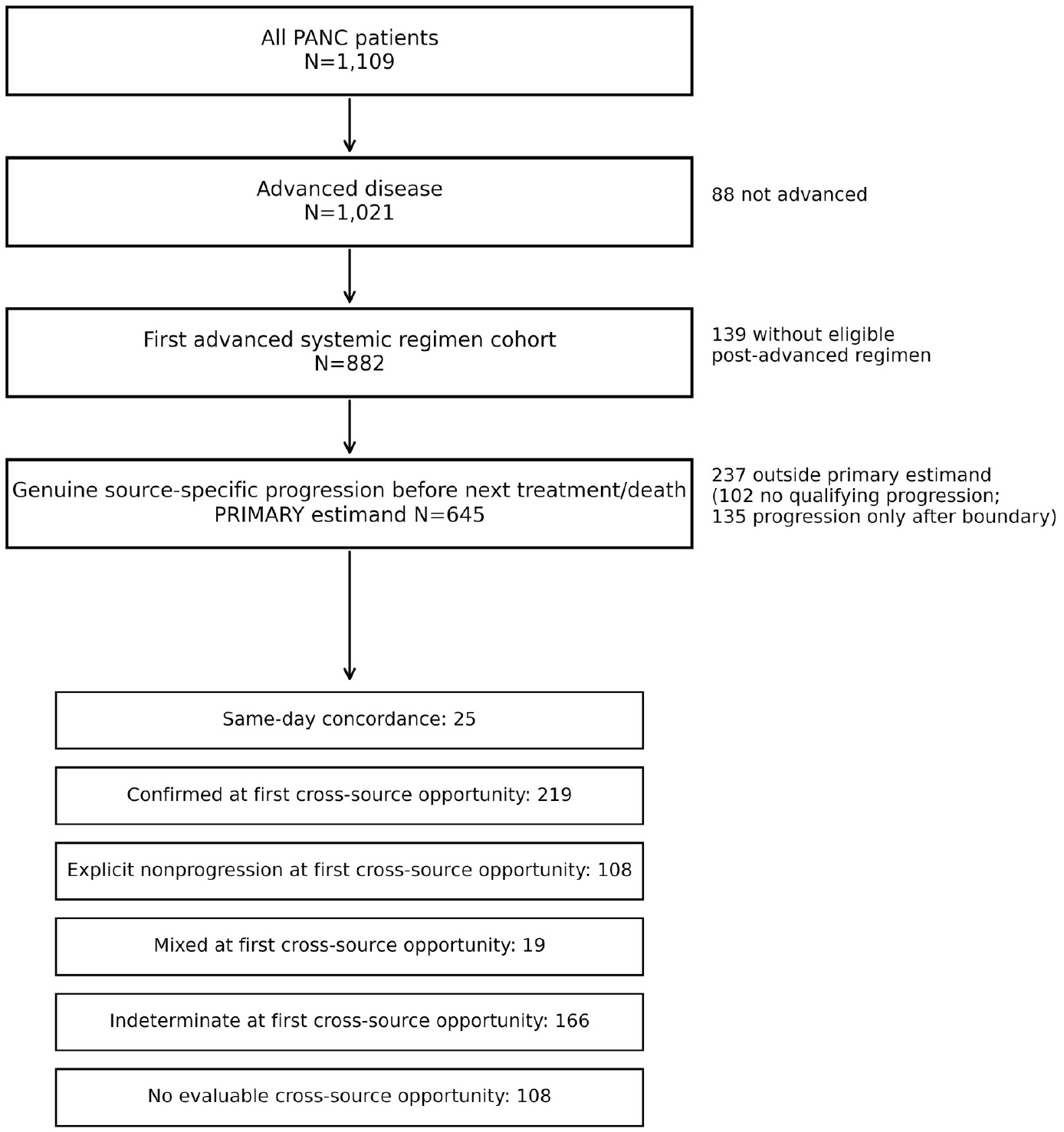
Cohort and endpoint flow. Flow from all PANC patients to advanced disease, the first advanced-regimen cohort, the progression-process primary estimand, and all six primary endpoint states.

In the primary six-state distribution (N=645), 25 patients (3.88%; 95% CI, 2.64%-5.66%) had same-day concordance, 219 (33.95%; 95% CI, 30.40%-37.69%) were confirmed at the first cross-source opportunity, 108 (16.74%; 95% CI, 14.06%-19.82%) had explicit nonprogression at that first opportunity, 19 (2.95%; 95% CI, 1.89%-4.55%) were mixed, 166 (25.74%; 95% CI, 22.51%-29.25%) were indeterminate, and 108 (16.74%; 95% CI, 14.06%-19.82%) had no evaluable cross-source opportunity (Table 2; Figure 2). No state was removed from the primary denominator.

**Table 2.**

| Hierarchy | Measure | n | Denominator | Estimate | 95% CI | Timing / additional detail |
| --- | --- | --- | --- | --- | --- | --- |
| PRIMARY | Same-day concordance | 25 | 645 | 3.88% | 2.64%–5.66% |  |
| PRIMARY | Confirmed at first cross-source opportunity | 219 | 645 | 33.95% | 30.40%–37.69% |  |
| PRIMARY | Explicit nonprogression at first cross-source opportunity | 108 | 645 | 16.74% | 14.06%–19.82% |  |
| PRIMARY | Mixed at first cross-source opportunity | 19 | 645 | 2.95% | 1.89%–4.55% |  |
| PRIMARY | Indeterminate at first cross-source opportunity | 166 | 645 | 25.74% | 22.51%–29.25% |  |
| PRIMARY | No evaluable cross-source opportunity | 108 | 645 | 16.74% | 14.06%–19.82% |  |
| SECONDARY | Explicit nonprogression among progression/nonprogression-classifiable first opportunities | 108 | 327 | 33.03% | 28.15%–38.30% | Conditional estimate; selected by observation opportunity |
| SECONDARY_TIMING | T0 to first cross-source opportunity | 537 | 645 | — | — | Median 6 days (IQR 2–19) |

**Figure 2.**
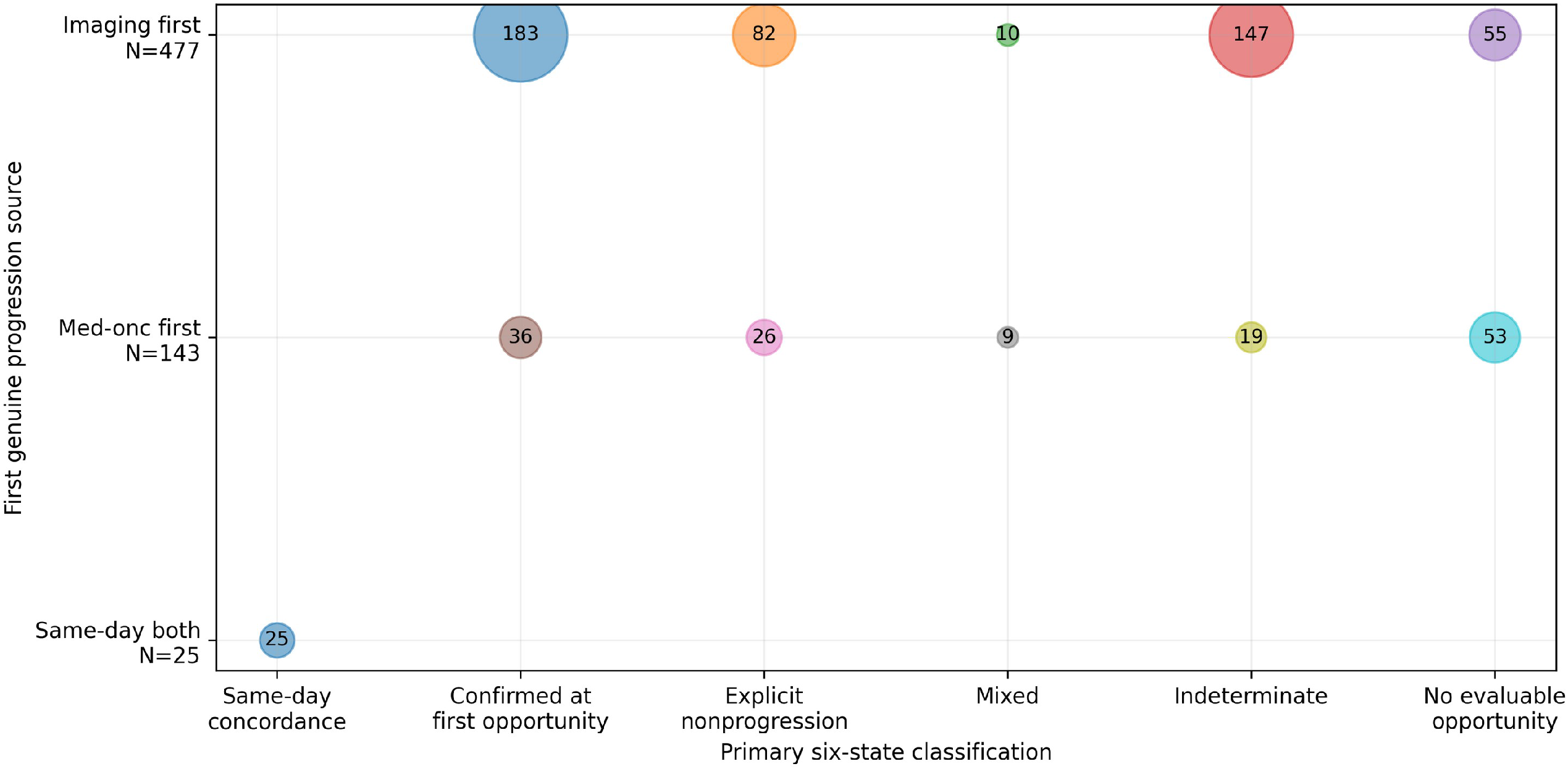
Source-to-state transition matrix. Bubble area is proportional to patient count for each first-progression-source × primary-state cell. This matrix is used as the clearest equivalent of an alluvial/Sankey display; no state is omitted.

### Secondary opportunity-conditioned estimate and timing

Among 327 first opportunities classifiable specifically as progression or explicit nonprogression, 108 (33.03%; 95% CI, 28.15%-38.30%) documented explicit nonprogression. This estimate applies only to patients with a classifiable opposite-source opportunity. Overall, 537 of 645 patients had a first opposite-source opportunity, at a median of 6 days after T0 (IQR, 2-19). Opposite-source progression confirmation was observed in 364 patients at a median of 13.5 days (IQR, 4-79); the Aalen-Johansen cumulative incidence of confirmation was 29.69% at 14 days, 34.88% at 30 days, 41.15% at 60 days, and 45.11% at 90 days.

### Unequal source-observation opportunity

Observation opportunity differed by index source (Figure 3). After imaging-first progression, 422 of 477 patients (88.47%) had a medical-oncology opportunity at a median of 5 days (IQR, 3-12). After medical-oncology-first progression, 90 of 143 (62.94%) had an imaging opportunity at a median of 38 days (IQR, 10.25-58.5). The source-standardized conditional explicit-nonprogression estimate was 33.48% and the measured-covariate inverse-probability-weighted diagnostic was 34.01%, close to the observed 33.03%. In contrast, deliberately extreme assignments of nonclassifiable patients spanned approximately 17.42%-64.68%, demonstrating nonidentification outside observed classifiable opportunities.

**Figure 3.**
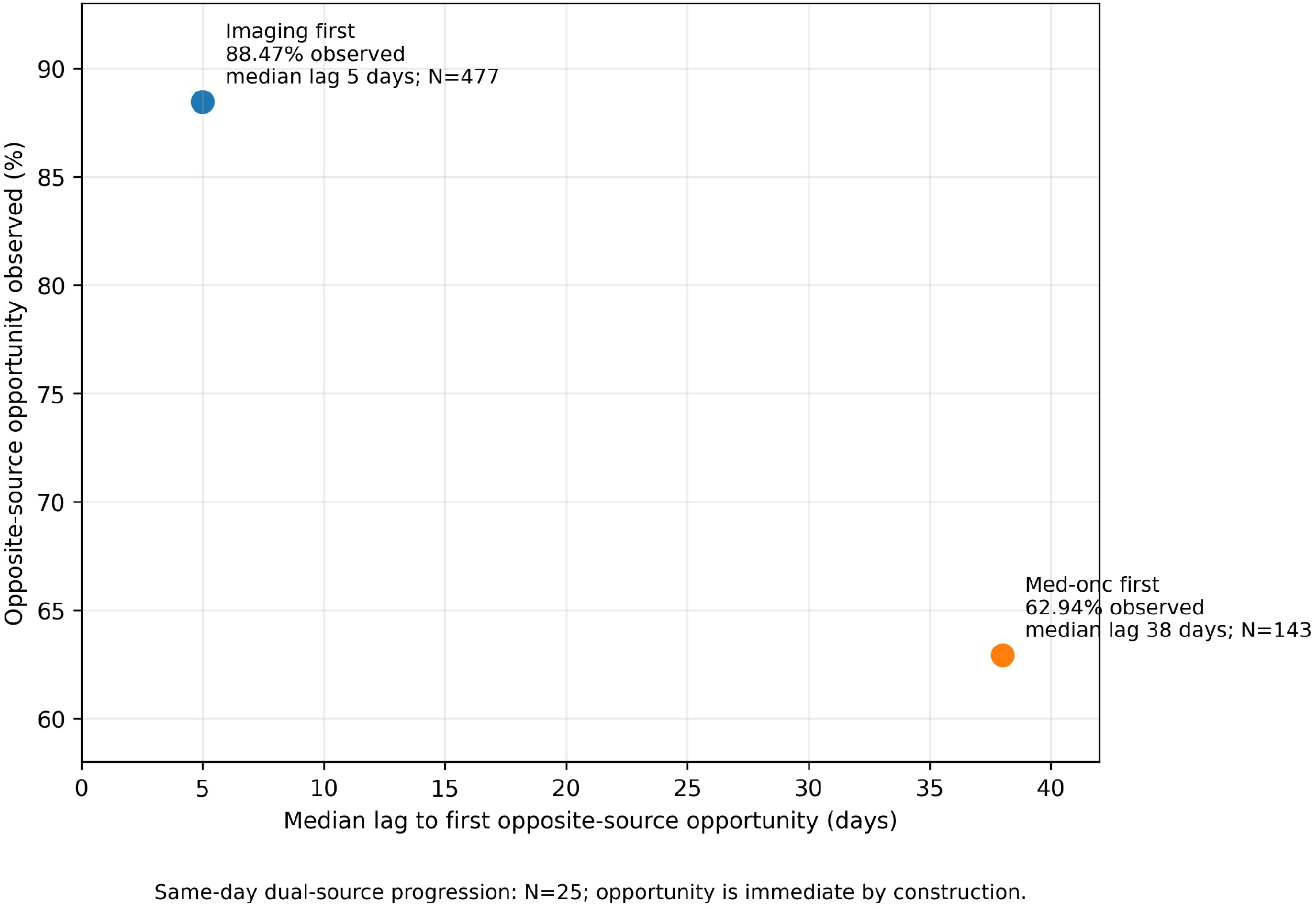
Unequal source-observation opportunity. Imaging-first progression has an opposite-source opportunity observed in 88.47% with median lag 5 days; medical-oncology-first progression has 62.94% observed with median lag 38 days. Same-day dual-source progression (N=25) is immediate by construction. The asymmetry is a structural feature of the measurement process and constrains interpretation of the conditional 108/327 estimate.

### Clinical context

An actual next same-cancer regimen was observed strictly after T0 in 443 of 645 patients (68.68%) and on T0 in 18 (2.79%); 461 patients (71.47%) therefore had a recorded next regimen on or after T0. Among those 461 patients, the median interval to the next regimen was 31 days (IQR, 11-112); among the 443 strictly-after-T0 transitions, the median was 36 days (IQR, 13-115). Because next-regimen timing participates in the endpoint boundary, these treatment-transition findings are descriptive and structurally linked rather than independent validation. Paired CA19-9 values in the prespecified ±60-day windows were available for 328 patients (50.9%); the median log1p(post) - log1p(pre) change was 0.118 (IQR, -0.114 to 0.471). Descriptive post-T0 median overall survival was 261 days (8.57 months), and 1-year survival was 37.77% (Table 3).

**Table 3.**
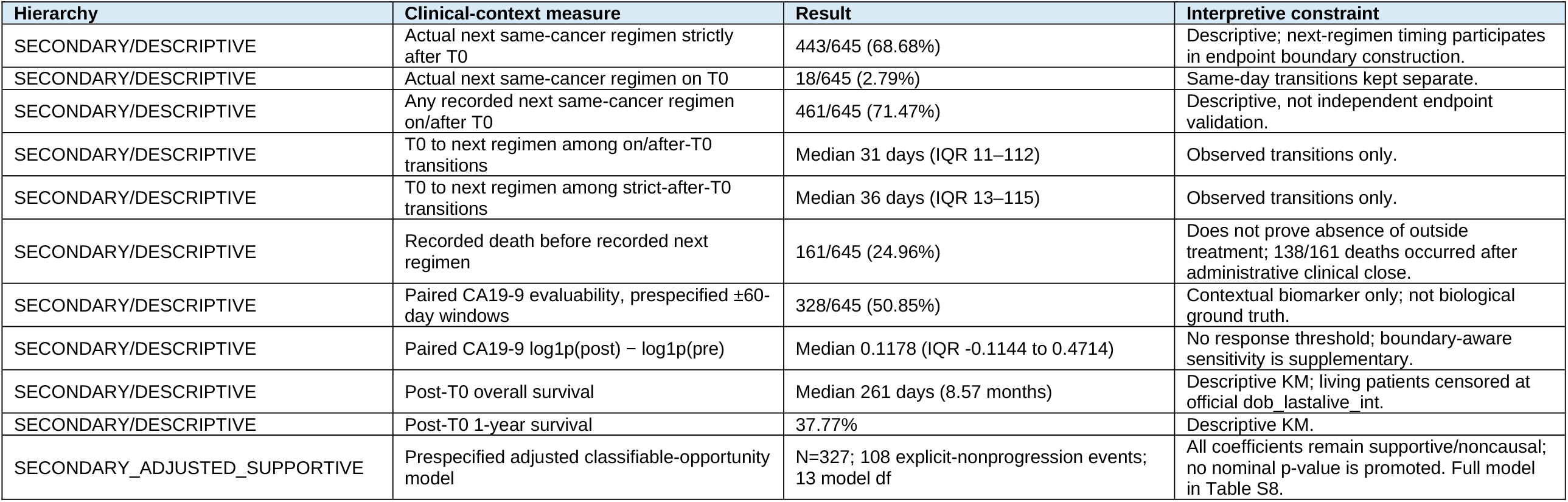

### Sensitivity and bias analyses

The ordinary endpoint-rule stress tests produced conditional explicit-nonprogression estimates close to the prespecified secondary estimate: 31.01% with strict same-day conflict handling, 31.21% when MIXED counted as confirmation, 30.00% with permissive boundary-day ordering, 33.42% with stricter cross-source evaluability, and 34.22% in the single-primary-cancer restriction (Table S3; Figure S2). The materially different sensitivity was the known-regimen-completion boundary, which reduced the estimand from 645 to 299 and yielded 84 explicit-nonprogression states among 144 classifiable opportunities (58.33%; 95% CI, 50.17%-66.07%). Among 71 patients missing dx_reg_end_all_int, 59 were recorded as discontinued, 7 as not discontinued, and 5 as unknown; 40 were maximally potentially unresolved with respect to completion timing. Excluding those 40 in an extreme stress exercise yielded N=259 and 81/120 explicit nonprogression (67.50%); this was treated as a stress bound, not a preferred analysis. The boundary-aware CA19-9 sensitivity retained 263 paired patients and produced a median log1p change of 0.108, close to the prespecified ±60-day estimate of 0.118 (Table S6). Descriptive heterogeneity was observed across presentation, institution, regimen, and observation-density strata, but these analyses were not interpreted as causal effect modification.

## Discussion

The central finding was the full six-state progression measurement process, not a single discordance percentage. Among 645 patients with qualifying source-specific progression, 33.9% were confirmed at the first opposite-source opportunity and 16.7% had explicit nonprogression, while 25.7% were indeterminate and 16.7% had no evaluable opposite-source opportunity. Within the narrower set of 327 classifiable progression/nonprogression opportunities, 33.0% showed explicit nonprogression. That conditional proportion is not a prevalence estimate of biological disagreement because entry into its denominator depended on the observation process. Retaining mixed, indeterminate, and no-opportunity states in the primary estimand avoids making classifiability a hidden eligibility criterion. [8-9,11]

The analysis builds on established radiology-anchored and clinician-anchored real-world progression methods rather than claiming to originate them. Prior work has compared the two approaches directly and has defined imaging-derived, medical-oncology-derived, and combined GENIE BPC progression endpoints. [5,4,3] The present contribution is narrower: the first documented progression from either source defines T0, the opposite source is evaluated at its first eligible opportunity, and the resulting state space retains indeterminate and unobserved opportunities instead of collapsing them into a binary comparison.

Observation opportunity was strongly source-dependent. After imaging-first progression, 88.5% of patients had a medical-oncology opportunity at a median of 5 days; after medical-oncology-first progression, only 62.9% had an imaging opportunity and the median lag was 38 days. This asymmetry accords with evidence that measured progression depends on assessment schedules, intermittent observation, and differential imaging frequency. [8-11] It does not establish why any individual pair of records differed, but it shows that radiology and medical-oncology documentation operated on non-equivalent observation clocks. Similar observed, source-standardized, and measured-covariate-weighted conditional estimates reduce concern about measured selection alone, whereas the wide extreme nonidentification bounds show that unobserved or nonclassifiable opportunities cannot simply be assigned a benign interpretation.

Clinical-boundary choice produced the largest quantitative shift. Adding documented completion of all drugs in the index regimen to the next-treatment/death boundary reduced the estimand from 645 to 299 and increased the conditional explicit-nonprogression proportion from 33.0% to 58.3%. The PANC definition of dx_reg_end_all_int supports this as a known-regimen-completion sensitivity, not a complete on-treatment analysis. [2] Regimen-end timing was missing for 71 patients, including 40 maximally unresolved in the timing stress set; excluding those 40 yielded a conditional proportion of 67.5%. The finding therefore persisted while its magnitude changed substantially with the boundary. The primary population should consequently be described as progression after initiation of the first advanced systemic regimen and before subsequent same-cancer treatment or death, rather than as progression while on first-line therapy.

The ancillary clinical-context analyses should also remain subordinate. CA19-9 can be useful for longitudinal pancreatic cancer monitoring, but guidelines do not support substituting it for imaging, and its interpretation is limited by biological and clinical factors. [16,19-20] The similar median CA19-9 changes across prespecified window and boundary-aware sensitivities argue against the contextual trajectory being driven solely by a convenient time window, but CA19-9 cannot identify which source was biologically correct. Likewise, subsequent treatment was common after T0, yet treatment-transition-by-state is structurally linked to the endpoint because next-regimen timing helps terminate the cross-source opportunity window. It therefore provides clinical description rather than independent validation.

The novelty claim is deliberately narrow. Contemporary PDAC studies have addressed treatment decisions after RECIST-defined progression, prediction of imminent progression from serial CT, and genomic outcomes in the same dataset family. [12-14] None located in the prespecified search through September 9, 2026 reproduced the complete source-first, first-opposite-source-opportunity six-state process used here. Radiology-versus-clinician progression and PFS-I/PFS-M remain established prior art; the distinct contribution is the integrated pancreatic-specific state-space, opportunity, timing, and boundary design. [5,4]

Strengths include separately curated longitudinal source fields, a prespecified source-first T0, an exhaustive outcome that retained nonclassifiable and unobserved opportunities, patient-level traceability, and extensive sensitivity analyses. Important limitations remain. Assessment timing was uneven and source-dependent; curated source labels were not a centralized RECIST rereview; treatment and outside-care capture may be incomplete after administrative observation ends; regimen-completion timing was missing for some patients; and neither CA19-9 nor subsequent treatment provides a biological gold standard. The analysis is descriptive and process-focused rather than causal, and heterogeneity across institutions, regimens, and observation density should not be interpreted as effect modification. These limitations are consistent with broader concerns about progression measurement under intermittent and differential observation in real-world oncology data. [7-9,11]

Cross-source progression assessment in advanced pancreatic cancer could not be reduced reliably to a single binary discordance estimate. Explicit nonprogression, indeterminate assessments, and absent opposite-source opportunities all contributed materially, and the estimated magnitude changed with both observation opportunity and clinical-boundary definition. The six-state framework makes those dependencies explicit. In the literature searched through September 9, 2026, this integrated pancreatic-specific source-first/first-opposite-source-opportunity approach remained distinct from established radiology-versus-clinician and GENIE source-specific progression methods. [5,4]

## Data Availability

The source dataset, AACR Project GENIE BPC PANC v1.0-public, is available through AACR Project GENIE under its applicable access and use terms. The authors do not redistribute GENIE source-level data. Reproducibility code that does not embed GENIE source data or patient-level derived records is available from the corresponding author on reasonable request, subject to applicable GENIE terms.

https://aacrprojectgenie.org/data/panc-1-0-public/

## Acknowledgments

The authors acknowledge the American Association for Cancer Research for its support in developing the AACR Project GENIE registry and the consortium members for their commitment to data sharing. Interpretations are the responsibility of the study authors.

OpenAI GPT-5.6 Sol (OpenAI; accessed September 9, 2026) also assisted with manuscript drafting, structural editing, and journal-format adaptation; its research-workflow use is described in Methods. The authors reviewed all outputs and retain full responsibility for the submitted work.

## Author contributions

Conception and design: All authors. Collection and assembly of data: All authors. Data analysis and interpretation: All authors. Manuscript writing: All authors. Final approval of manuscript: All authors.

