## Supplementary material for "Cross-Source Progression Assessment in Advanced Pancreatic Cancer: An Opportunity-Adjusted Six-State Analysis": Tables S1-S8 and Figures S1-S5

Contents: Tables S1–S8 and Figures S1–S5. Wide tables are split into continued column panels solely for readability; all rows, columns, and values are preserved from the source display files.

#### Table S1. Definitions and estimands

##### All columns

| Type | Term | Count/Denominator | Operational definition / interpretation |
|---|---|---|---|
| Primary cohort | Progression-process cohort | 645 | First genuine source-specific progression after first advanced-regimen initiation and not after the next same-cancer treatment/death boundary. |
| PRIMARY state | Same-day concordance | 25 | Both sources first show genuine progression on T0. |
| PRIMARY state | Confirmed at first opportunity | 219 | Opposite source first evaluable assessment shows progression. |
| PRIMARY state | Explicit nonprogression | 108 | Opposite source first evaluable assessment explicitly shows stable/no change or improving/responding. |
| PRIMARY state | Mixed | 19 | Opposite source first evaluable assessment is mixed. |
| PRIMARY state | Indeterminate | 166 | First opportunity cannot be classified as progression/nonprogression/mixed under prespecified rules, including boundary-day uncertainty. |
| PRIMARY state | No evaluable opportunity | 108 | No opposite-source assessment before the prespecified clinical boundary or administrative close. |
| SECONDARY estimand | Conditional explicit nonprogression | 108/327 | Explicit nonprogression divided by confirmed + explicit nonprogression only; selected by observation opportunity. |
| Boundary sensitivity | Known-regimen-completion boundary | 299 | Adds known end of all drugs in the index regimen (dx_reg_end_all_int) when documented; not a complete on-treatment analysis. |

### Table S2. Evaluability and source-observation process

#### Continued panel 1 of 2

| analysis_id | stratum | n | opportunity_observed_n | opportunity_observed_percent | classifiable_first_opportunity_n | classifiable_first_opportunity_percent |
|---|---|---|---|---|---|---|
| INDEX_SOURCE | IMAGING_FIRST | 477 | 422 | 88.46960168 | 265 | 55.55555556 |
| INDEX_SOURCE | MEDONC_FIRST | 143 | 90 | 62.93706294 | 62 | 43.35664336 |
| INDEX_SOURCE | SAME_DAY_BOTH | 25 | 25 | 100.0 | 0 | 0.0 |
| INSTITUTION | DFCI | 245 | 204 | 83.26530612 | 128 | 52.24489796 |
| INSTITUTION | MSK | 309 | 260 | 84.14239482 | 153 | 49.51456311 |
| INSTITUTION | UHN | 46 | 35 | 76.08695652 | 27 | 58.69565217 |
| INSTITUTION | VICC | 45 | 38 | 84.44444444 | 19 | 42.22222222 |
| PRESENTATION | De novo Stage IV | 323 | 266 | 82.35294118 | 156 | 48.29721362 |
| PRESENTATION | Stage I-III then distant mets | 322 | 271 | 84.16149068 | 171 | 53.10559006 |
| REGIMEN_CLASS | FOLFIRINOX/mFOLFIRINOX-containing | 277 | 223 | 80.50541516 | 145 | 52.3465704 |
| REGIMEN_CLASS | Fluoropyrimidine/oxaliplatin-based | 28 | 25 | 89.28571429 | 14 | 50.0 |
| REGIMEN_CLASS | Gemcitabine+nab-paclitaxel | 175 | 156 | 89.14285714 | 87 | 49.71428571 |
| REGIMEN_CLASS | Other | 54 | 45 | 83.33333333 | 30 | 55.55555556 |
| REGIMEN_CLASS | Other gemcitabine-based | 111 | 88 | 79.27927928 | 51 | 45.94594595 |
| OBSERVATION_DENSITY_COUNT_QUARTILE_APPROX | LOW_0_1 | 208 | 176 | 84.61538462 | 106 | 50.96153846 |
| OBSERVATION_DENSITY_COUNT_QUARTILE_APPROX | LOWMID_2_3 | 124 | 103 | 83.06451613 | 60 | 48.38709677 |
| OBSERVATION_DENSITY_COUNT_QUARTILE_APPROX | HIGHMID_4_8 | 165 | 126 | 76.36363636 | 81 | 49.09090909 |
| OBSERVATION_DENSITY_COUNT_QUARTILE_APPROX | HIGH_9PLUS | 148 | 132 | 89.18918919 | 80 | 54.05405405 |

#### Continued panel 2 of 2

| analysis_id | stratum | indeterminate_or_noopportunity_n | indeterminate_or_noopportunity_percent | first_opportunity_lag_median_days | first_opportunity_lag_q1_days | first_opportunity_lag_q3_days |
|---|---|---|---|---|---|---|
| INDEX_SOURCE | IMAGING_FIRST | 202 | 42.34800839 | 5.0 | 3.0 | 12.0 |
| INDEX_SOURCE | MEDONC_FIRST | 72 | 50.34965035 | 38.0 | 10.25 | 58.5 |
| INDEX_SOURCE | SAME_DAY_BOTH | 0 | 0.0 | 0.0 | 0.0 | 0.0 |
| INSTITUTION | DFCI | 98 | 40.0 | 6.0 | 2.0 | 17.5 |
| INSTITUTION | MSK | 136 | 44.01294498 | 5.0 | 2.0 | 18.0 |
| INSTITUTION | UHN | 19 | 41.30434783 | 8.0 | 6.0 | 13.0 |
| INSTITUTION | VICC | 21 | 46.66666667 | 6.0 | 4.0 | 36.5 |
| PRESENTATION | De novo Stage IV | 143 | 44.27244582 | 5.0 | 1.25 | 13.75 |
| PRESENTATION | Stage I-III then distant mets | 131 | 40.68322981 | 7.0 | 3.0 | 24.5 |
| REGIMEN_CLASS | FOLFIRINOX/mFOLFIRINOX-containing | 118 | 42.59927798 | 5.0 | 2.0 | 12.0 |
| REGIMEN_CLASS | Fluoropyrimidine/oxaliplatin-based | 10 | 35.71428571 | 8.0 | 2.0 | 19.0 |
| REGIMEN_CLASS | Gemcitabine+nab-paclitaxel | 69 | 39.42857143 | 6.0 | 3.0 | 21.25 |
| REGIMEN_CLASS | Other | 23 | 42.59259259 | 7.0 | 3.0 | 29.0 |
| REGIMEN_CLASS | Other gemcitabine-based | 54 | 48.64864865 | 6.5 | 3.0 | 23.75 |
| OBSERVATION_DENSITY_COUNT_QUARTILE_APPROX | LOW_0_1 | 88 | 42.30769231 | 11.0 | 4.0 | 34.25 |
| OBSERVATION_DENSITY_COUNT_QUARTILE_APPROX | LOWMID_2_3 | 55 | 44.35483871 | 5.0 | 2.0 | 9.5 |
| OBSERVATION_DENSITY_COUNT_QUARTILE_APPROX | HIGHMID_4_8 | 72 | 43.63636364 | 5.0 | 2.0 | 11.75 |
| OBSERVATION_DENSITY_COUNT_QUARTILE_APPROX | HIGH_9PLUS | 59 | 39.86486486 | 5.0 | 1.0 | 12.25 |

*Note: Imaging-first and med-onc-first opportunity processes occur on materially different clocks.*

### Table S3. Endpoint sensitivity battery

#### Continued panel 1 of 3

| sensitivity_id | estimand_n | same_day_n | confirmed_n | explicit_nonprogression_n | mixed_n | indeterminate_n |
|---|---|---|---|---|---|---|
| PRIMARY_REFERENCE | 645 | 25 | 219 | 108 | 19 | 166 |
| STRICT_SAME_DAY_CONFLICT | 638 | 25 | 218 | 98 | 18 | 168 |
| MIXED_AS_PROGRESSION | 645 | 25 | 238 | 108 | 0 | 166 |
| PERMISSIVE_BOUNDARY_DAY | 645 | 25 | 259 | 111 | 20 | 122 |
| STRICT_CROSS_SOURCE_EVALUABILITY | 645 | 25 | 261 | 131 | 21 | 47 |
| ALT_BOUNDARY_INDEX_REGIMEN_END_ALL | 299 | 9 | 60 | 84 | 11 | 78 |
| SINGLE_PRIMARY_CANCER | 516 | 22 | 173 | 90 | 16 | 129 |

#### Continued panel 2 of 3

| sensitivity_id | no_evaluable_opportunity_n | explicit_nonprogression_percent_all | classifiable_n | conditional_explicit_nonprogression_percent | conditional_ci95_low_percent | conditional_ci95_high_percent |
|---|---|---|---|---|---|---|
| PRIMARY_REFERENCE | 108 | 16.74418605 | 327 | 33.02752294 | 28.15290664 | 38.29628011 |
| STRICT_SAME_DAY_CONFLICT | 111 | 15.36050157 | 316 | 31.01265823 | 26.1664315 | 36.31498025 |
| MIXED_AS_PROGRESSION | 108 | 16.74418605 | 346 | 31.21387283 | 26.56024458 | 36.28006588 |
| PERMISSIVE_BOUNDARY_DAY | 108 | 17.20930233 | 370 | 30.0 | 25.55566633 | 34.85535911 |
| STRICT_CROSS_SOURCE_EVALUABILITY | 160 | 20.31007752 | 392 | 33.41836735 | 28.92966368 | 38.22890521 |
| ALT_BOUNDARY_INDEX_REGIMEN_END_ALL | 57 | 28.09364548 | 144 | 58.33333333 | 50.16686221 | 66.06674384 |
| SINGLE_PRIMARY_CANCER | 86 | 17.44186047 | 263 | 34.22053232 | 28.75057228 | 40.14481588 |

#### Continued panel 3 of 3

| sensitivity_id | patient_assignments_changed_vs_primary | primary_estimand_to_nonestimand_n | nonestimand_to_primary_estimand_n | notes | conditional_nonprogression_delta_pp_vs_primary |
|---|---|---|---|---|---|
| PRIMARY_REFERENCE | | | | Primary endpoint reconstruction. | 0.0 |
| STRICT_SAME_DAY_CONFLICT | 22.0 | 7.0 | 0.0 | Imaging patient-days containing both progression and explicit nonprogression map to INDETERMINATE rather than progression-priority. | -2.014864708 |
| MIXED_AS_PROGRESSION | 19.0 | 0.0 | 0.0 | Explicit MIXED at the opposite-source first opportunity is counted as confirmation; T0 definition is unchanged. | -1.813650103 |
| PERMISSIVE_BOUNDARY_DAY | 44.0 | 0.0 | 0.0 | Opposite-source assessment on the same date as treatment/death boundary is classified by observed state rather than forced INDETERMINATE. | -3.027522936 |
| STRICT_CROSS_SOURCE_EVALUABILITY | 119.0 | 0.0 | 0.0 | Opposite-source opportunity requires curated PROGRESSION/NONPROGRESSIVE/MIXED content; indeterminate-only days are skipped. | 0.3908444112 |
| ALT_BOUNDARY_INDEX_REGIMEN_END_ALL | 387.0 | 346.0 | 0.0 | Clinical boundary is min(next same-cancer regimen, death, known end of all drugs in index regimen); only known-discontinued regimen end-all dates are used. | 25.3058104 |
| SINGLE_PRIMARY_CANCER | | | | Primary endpoint assignments restricted to patients with n_cancers == 1. | 1.193009384 |

*Note: The alternate boundary is a known-regimen-completion sensitivity using dx_reg_end_all_int only when documented.*

*Note: It must not be described as a complete on-treatment analysis because regimen-end information is incompletely observed.*

### Table S4. Descriptive heterogeneity by presentation, institution, regimen, and observation density

#### Continued panel 1 of 3

| analysis_id | stratum | n | same_day_n | confirmed_n | explicit_nonprogression_n | mixed_n | indeterminate_n |
|---|---|---|---|---|---|---|---|
| PRESENTATION | De novo Stage IV | 323 | 12 | 94 | 62 | 12 | 86 |
| PRESENTATION | Stage I-III then distant mets | 322 | 13 | 125 | 46 | 7 | 80 |
| INSTITUTION | DFCI | 245 | 13 | 80 | 48 | 6 | 57 |
| INSTITUTION | MSK | 309 | 9 | 107 | 46 | 11 | 87 |
| INSTITUTION | UHN | 46 | 0 | 21 | 6 | 0 | 8 |
| INSTITUTION | VICC | 45 | 3 | 11 | 8 | 2 | 14 |
| REGIMEN_CLASS | FOLFIRINOX/ mFOLFIRINOX-containing | 277 | 6 | 94 | 51 | 8 | 64 |
| REGIMEN_CLASS | Fluoropyrimidine/oxaliplatin-based | 28 | 3 | 10 | 4 | 1 | 7 |
| REGIMEN_CLASS | Gemcitabine+nab-paclitaxel | 175 | 11 | 48 | 39 | 8 | 50 |
| REGIMEN_CLASS | Other | 54 | 1 | 23 | 7 | 0 | 14 |
| REGIMEN_CLASS | Other gemcitabine-based | 111 | 4 | 44 | 7 | 2 | 31 |
| OBSERVATION_DENSITY_COUNT_QUARTILE_APPROX | LOW_0_1 | 208 | 7 | 63 | 43 | 7 | 56 |
| OBSERVATION_DENSITY_COUNT_QUARTILE_APPROX | LOWMID_2_3 | 124 | 5 | 39 | 21 | 4 | 34 |
| OBSERVATION_DENSITY_COUNT_QUARTILE_APPROX | HIGHMID_4_8 | 165 | 5 | 55 | 26 | 7 | 33 |
| OBSERVATION_DENSITY_COUNT_QUARTILE_APPROX | HIGH_9PLUS | 148 | 8 | 62 | 18 | 1 | 43 |

#### Continued panel 2 of 3

| analysis_id | stratum | no_evaluable_opportunity_n | confirmed_percent | explicit_nonprogression_percent_all | indeterminate_percent | no_evaluable_opportunity_percent | classifiable_n |
|---|---|---|---|---|---|---|---|
| PRESENTATION | De novo Stage IV | 57 | 29.10216718 | 19.19504644 | 26.625387 | 17.64705882 | 156 |
| PRESENTATION | Stage I-III then distant mets | 51 | 38.81987578 | 14.28571429 | 24.8447205 | 15.83850932 | 171 |
| INSTITUTION | DFCI | 41 | 32.65306122 | 19.59183673 | 23.26530612 | 16.73469388 | 128 |
| INSTITUTION | MSK | 49 | 34.62783172 | 14.88673139 | 28.15533981 | 15.85760518 | 153 |
| INSTITUTION | UHN | 11 | 45.65217391 | 13.04347826 | 17.39130435 | 23.91304348 | 27 |
| INSTITUTION | VICC | 7 | 24.44444444 | 17.77777778 | 31.11111111 | 15.55555556 | 19 |
| REGIMEN_CLASS | FOLFIRINOX/ mFOLFIRINOX-containing | 54 | 33.93501805 | 18.41155235 | 23.10469314 | 19.49458484 | 145 |
| REGIMEN_CLASS | Fluoropyrimidine/oxaliplatin-based | 3 | 35.71428571 | 14.28571429 | 25.0 | 10.71428571 | 14 |
| REGIMEN_CLASS | Gemcitabine+nab-paclitaxel | 19 | 27.42857143 | 22.28571429 | 28.57142857 | 10.85714286 | 87 |
| REGIMEN_CLASS | Other | 9 | 42.59259259 | 12.96296296 | 25.92592593 | 16.66666667 | 30 |
| REGIMEN_CLASS | Other gemcitabine-based | 23 | 39.63963964 | 6.306306306 | 27.92792793 | 20.72072072 | 51 |
| OBSERVATION_DENSITY_COUNT_QUARTILE_APPROX | LOW_0_1 | 32 | 30.28846154 | 20.67307692 | 26.92307692 | 15.38461538 | 106 |
| OBSERVATION_DENSITY_COUNT_QUARTILE_APPROX | LOWMID_2_3 | 21 | 31.4516129 | 16.93548387 | 27.41935484 | 16.93548387 | 60 |
| OBSERVATION_DENSITY_COUNT_QUARTILE_APPROX | HIGHMID_4_8 | 39 | 33.33333333 | 15.75757576 | 20.0 | 23.63636364 | 81 |
| OBSERVATION_DENSITY_COUNT_QUARTILE_APPROX | HIGH_9PLUS | 16 | 41.89189189 | 12.16216216 | 29.05405405 | 10.81081081 | 80 |

#### Continued panel 3 of 3

| analysis_id | stratum | conditional_explicit_nonprogression_percent | conditional_ci95_low_percent | conditional_ci95_high_percent |
|---|---|---|---|---|
| PRESENTATION | De novo Stage IV | 39.74358974 | 32.39962894 | 47.58053375 |
| PRESENTATION | Stage I-III then distant mets | 26.9005848 | 20.81553903 | 34.00066959 |
| INSTITUTION | DFCI | 37.5 | 29.5924291 | 46.13599481 |
| INSTITUTION | MSK | 30.06535948 | 23.36077944 | 37.74644281 |
| INSTITUTION | UHN | 22.22222222 | 10.60723945 | 40.75692887 |
| INSTITUTION | VICC | 42.10526316 | 23.14189128 | 63.72409647 |

| analysis_id | stratum | conditional_explicit_nonprogression_percent | conditional_ci95_low_percent | conditional_ci95_high_percent |
|---|---|---|---|---|
| REGIMEN_CLASS | FOLFIRINOX/mFOLFIRINOX-containing | 35.17241379 | 27.87429277 | 43.23590707 |
| REGIMEN_CLASS | Fluoropyrimidine/oxaliplatin-based | 28.57142857 | 11.72137864 | 54.64908433 |
| REGIMEN_CLASS | Gemcitabine+nab-paclitaxel | 44.82758621 | 34.81717713 | 55.27545225 |
| REGIMEN_CLASS | Other | 23.33333333 | 11.79238814 | 40.92832616 |
| REGIMEN_CLASS | Other gemcitabine-based | 13.7254902 | 6.811101735 | 25.72169226 |
| OBSERVATION_DENSITY_COUNT_QUARTILE_APPROX | LOW_0_1 | 40.56603774 | 31.70748814 | 50.0844507 |
| OBSERVATION_DENSITY_COUNT_QUARTILE_APPROX | LOWMID_2_3 | 35.0 | 24.16777441 | 47.63738116 |
| OBSERVATION_DENSITY_COUNT_QUARTILE_APPROX | HIGHMID_4_8 | 32.09876543 | 22.94222197 | 42.87637615 |
| OBSERVATION_DENSITY_COUNT_QUARTILE_APPROX | HIGH_9PLUS | 22.5 | 14.73320734 | 32.78678978 |

*Note: These strata are descriptive/supportive; they are not causal effect-modifier analyses.*

### Table S5. Confirmation cumulative incidence at prespecified horizons

#### Continued panel 1 of 2

| sensitivity_id | analysis_label | time_horizon_days | method | confirmation_cif | confirmation_cif_percent | observed_confirmation_events_by_horizon |
|---|---|---|---|---|---|---|
| CONFIRMATION_WINDOW | SECONDARY_TIMING | 14 | AALEN_JOHANSEN_CIF | 0.2968626782 | 29.68626782 | 188 |
| CONFIRMATION_WINDOW | SECONDARY_TIMING | 30 | AALEN_JOHANSEN_CIF | 0.3487666633 | 34.87666633 | 220 |
| CONFIRMATION_WINDOW | SECONDARY_TIMING | 60 | AALEN_JOHANSEN_CIF | 0.4115374691 | 41.15374691 | 257 |
| CONFIRMATION_WINDOW | SECONDARY_TIMING | 90 | AALEN_JOHANSEN_CIF | 0.4510547381 | 45.10547381 | 279 |

#### Continued panel 2 of 2

| sensitivity_id | analysis_label | observed_competing_boundaries_by_horizon | administrative_censors_by_horizon | starting_denominator | interpretation |
|---|---|---|---|---|---|
| CONFIRMATION_WINDOW | SECONDARY_TIMING | 116 | 17 | 645 | Prespecified competing-risk confirmation profile; no outcome-driven window selection. |
| CONFIRMATION_WINDOW | SECONDARY_TIMING | 140 | 26 | 645 | Prespecified competing-risk confirmation profile; no outcome-driven window selection. |
| CONFIRMATION_WINDOW | SECONDARY_TIMING | 158 | 38 | 645 | Prespecified competing-risk confirmation profile; no outcome-driven window selection. |
| CONFIRMATION_WINDOW | SECONDARY_TIMING | 170 | 47 | 645 | Prespecified competing-risk confirmation profile; no outcome-driven window selection. |

*Note: Aalen–Johansen cumulative incidence with treatment/death as competing events and administrative close as censoring.*

### Table S6. CA19-9 window and clinical-boundary sensitivities

#### Continued panel 1 of 2

| sensitivity_id | pre_window | post_window | pre_evaluable_n | post_evaluable_n | paired_n | raw_change_q1 | raw_change_median |
|---|---|---|---|---|---|---|---|
| CA19_WINDOW_30D | [-30,-1] | [0,+30] | 304.0 | 395.0 | 237.0 | -15.0 | 13.0 |
| CA19_WINDOW_60D | [-60,-1] | [0,+60] | 375.0 | 446.0 | 328.0 | -7.25 | 15.0 |
| CA19_WINDOW_90D | [-90,-1] | [0,+90] | 432.0 | 470.0 | 379.0 | -5.0 | 14.0 |
| STRICTLY_BEFORE_BOUNDARY | | | | | | -7.5 | 14.0 |
| NO_OBSERVED_BOUNDARY | | | | | | -3.0 | 2.5 |
| SAME_BOUNDARY_DAY_INDETERMINATE | | | | | | -1.0 | 126.0 |
| STRICTLY_AFTER_BOUNDARY_EXCLUDED | | | | | | -241.0 | 4.0 |
| BOUNDARY_AWARE_ELIGIBLE_COMBINED | | | | | | -7.5 | 12.0 |

#### Continued panel 2 of 2

| sensitivity_id | raw_change_q3 | log1p_change_q1 | log1p_change_median | log1p_change_q3 | notes | analysis_group | n |
|---|---|---|---|---|---|---|---|
| CA19_WINDOW_30D | 493.0 | -0.1269307086 | 0.08792189352 | 0.3764775712 | Closest pre and earliest post patient-day after median collapse of same-day duplicate values. | WINDOW_SENSITIVITY | |
| CA19_WINDOW_60D | 575.0 | -0.1144422729 | 0.1178038348 | 0.4713551129 | Closest pre and earliest post patient-day after median collapse of same-day duplicate values. | WINDOW_SENSITIVITY | |
| CA19_WINDOW_90D | 518.0 | -0.1155558605 | 0.1316205554 | 0.5243716573 | Closest pre and earliest post patient-day after median collapse of same-day duplicate values. | WINDOW_SENSITIVITY | |
| STRICTLY_BEFORE_BOUNDARY | 393.5 | -0.08004270767 | 0.1082781517 | 0.4490409076 | | BOUNDARY_AWARE | 259.0 |
| NO_OBSERVED_BOUNDARY | 81.5 | -0.04296256422 | 0.1952524043 | 0.4661654015 | | BOUNDARY_AWARE | 4.0 |
| SAME_BOUNDARY_DAY_INDETERMINATE | 1473.75 | -0.04518681321 | 0.2310873685 | 0.4645256627 | | BOUNDARY_AWARE | 44.0 |
| STRICTLY_AFTER_BOUNDARY_EXCLUDED | 1973.0 | -0.535961597 | 0.07320340402 | 0.5877866649 | | BOUNDARY_AWARE | 21.0 |
| BOUNDARY_AWARE_ELIGIBLE_COMBINED | 344.0 | -0.08004270767 | 0.1082781517 | 0.4490409076 | | BOUNDARY_AWARE | 263.0 |

*Note: CA19-9 remains contextual and must not adjudicate which clinical source was biologically correct.*

### Table S7. Treatment-transition and post-T0 survival context

#### All columns

| Domain | Measure | Result | Constraint |
|---|---|---|---|
| Treatment cumulative incidence | 14 days | 23.95% | Descriptive; treatment is structurally linked to endpoint boundary. |
| Treatment cumulative incidence | 30 days | 37.15% | Descriptive; treatment is structurally linked to endpoint boundary. |
| Treatment cumulative incidence | 60 days | 47.51% | Descriptive; treatment is structurally linked to endpoint boundary. |
| Treatment cumulative incidence | 90 days | 53.93% | Descriptive; treatment is structurally linked to endpoint boundary. |
| Post-T0 OS | POST_T0_OVERALL_SURVIVAL_KM | Median 261 days (8.57 months) | Descriptive KM; censoring uses dob_lastalive_int. |
| Post-T0 OS | POST_T0_OS_SURVIVAL_90D | 83.97% | Descriptive KM; censoring uses dob_lastalive_int. |
| Post-T0 OS | POST_T0_OS_SURVIVAL_180D | 65.17% | Descriptive KM; censoring uses dob_lastalive_int. |
| Post-T0 OS | POST_T0_OS_SURVIVAL_365D | 37.77% | Descriptive KM; censoring uses dob_lastalive_int. |
| Post-T0 OS | POST_T0_OS_SURVIVAL_730D | 16.76% | Descriptive KM; censoring uses dob_lastalive_int. |

*Note: Treatment transition is not independent endpoint validation.*

### Table S8. Prespecified adjusted classifiable-opportunity model

#### All columns

| analysis_label | term | beta | standard_error | odds_ratio | ci95_low | ci95_high | p_value_secondary |
|---|---|---|---|---|---|---|---|
| SECONDARY_ADJUSTED_SUPPORTIVE | Intercept | -0.7040876843 | 0.4052927155 | 0.494559563 | 0.223476993 | 1.094471328 | 0.08234613752 |
| SECONDARY_ADJUSTED_SUPPORTIVE | C(sex, Treatment(reference="Male"))[T.Female] | 0.3265245562 | 0.2569409064 | 1.386142288 | 0.837720835 | 2.29359276 | 0.2037941966 |
| SECONDARY_ADJUSTED_SUPPORTIVE | C(institution, Treatment(reference="MSK"))[T.DFCI] | 0.3294521766 | 0.271831635 | 1.390206332 | 0.8160105152 | 2.368442085 | 0.2255233154 |
| SECONDARY_ADJUSTED_SUPPORTIVE | C(institution, Treatment(reference="MSK"))[T.UHN] | -0.507216552 | 0.5339049456 | 0.6021693554 | 0.2114744658 | 1.71466532 | 0.34210573 |
| SECONDARY_ADJUSTED_SUPPORTIVE | C(institution, Treatment(reference="MSK"))[T.VICC] | 0.5218485375 | 0.5489981798 | 1.685139817 | 0.5745500177 | 4.942469957 | 0.3418344169 |
| SECONDARY_ADJUSTED_SUPPORTIVE | C(presentation, Treatment(reference="De novo Stage IV"))[T.Stage I-III then distant mets] | -0.6060502739 | 0.2897395642 | 0.5455012 | 0.3091497831 | 0.9625481741 | 0.0364647342 |
| SECONDARY_ADJUSTED_SUPPORTIVE | C(regimen_class, Treatment(reference="FOLFIRINOX/mFOLFIRINOX-containing"))[T.Fluoropyrimidine/oxaliplatin-based] | -0.2320982569 | 0.6710688187 | 0.7928682147 | 0.2128071393 | 2.95403626 | 0.7294452483 |
| SECONDARY_ADJUSTED_SUPPORTIVE | C(regimen_class, Treatment(reference="FOLFIRINOX/mFOLFIRINOX-containing"))[T.Gemcitabine+nab-paclitaxel] | 0.6272024671 | 0.3118791251 | 1.872365242 | 1.016058183 | 3.450345324 | 0.04432086906 |
| SECONDARY_ADJUSTED_SUPPORTIVE | C(regimen_class, Treatment(reference="FOLFIRINOX/mFOLFIRINOX-containing"))[T.Other] | -0.3439683784 | 0.516901916 | 0.7089513458 | 0.2574119607 | 1.952558884 | 0.5057676637 |
| SECONDARY_ADJUSTED_SUPPORTIVE | C(regimen_class, Treatment(reference="FOLFIRINOX/mFOLFIRINOX-containing"))[T.Other gemcitabine-based] | -0.817865717 | 0.4939029992 | 0.4413726641 | 0.1676464792 | 1.162027557 | 0.09773725705 |
| SECONDARY_ADJUSTED_SUPPORTIVE | C(index_source, Treatment(reference="IMAGING_FIRST"))[T.MEDONC_FIRST] | 0.6707398307 | 0.3525416583 | 1.95568366 | 0.9799741134 | 3.902856746 | 0.05709495509 |
| SECONDARY_ADJUSTED_SUPPORTIVE | age10 | -0.1849970707 | 0.1323581656 | 0.8311067184 | 0.6411997114 | 1.077259339 | 0.1622030321 |
| SECONDARY_ADJUSTED_SUPPORTIVE | log1p_imaging_pre_t0_count | -0.699695941 | 0.3389298943 | 0.496736318 | 0.2556396797 | 0.9652138897 | 0.03897727215 |
| SECONDARY_ADJUSTED_SUPPORTIVE | log1p_medonc_pre_t0_count | 0.261815138 | 0.3097519453 | 1.299286332 | 0.708016756 | 2.384329124 | 0.3979761256 |

*Note: All coefficients are SECONDARY/supportive/noncausal; nominal p-values do not determine prominence or model selection.*

#### Supplementary Figures

**Figure S1. Confirmation cumulative incidence.**

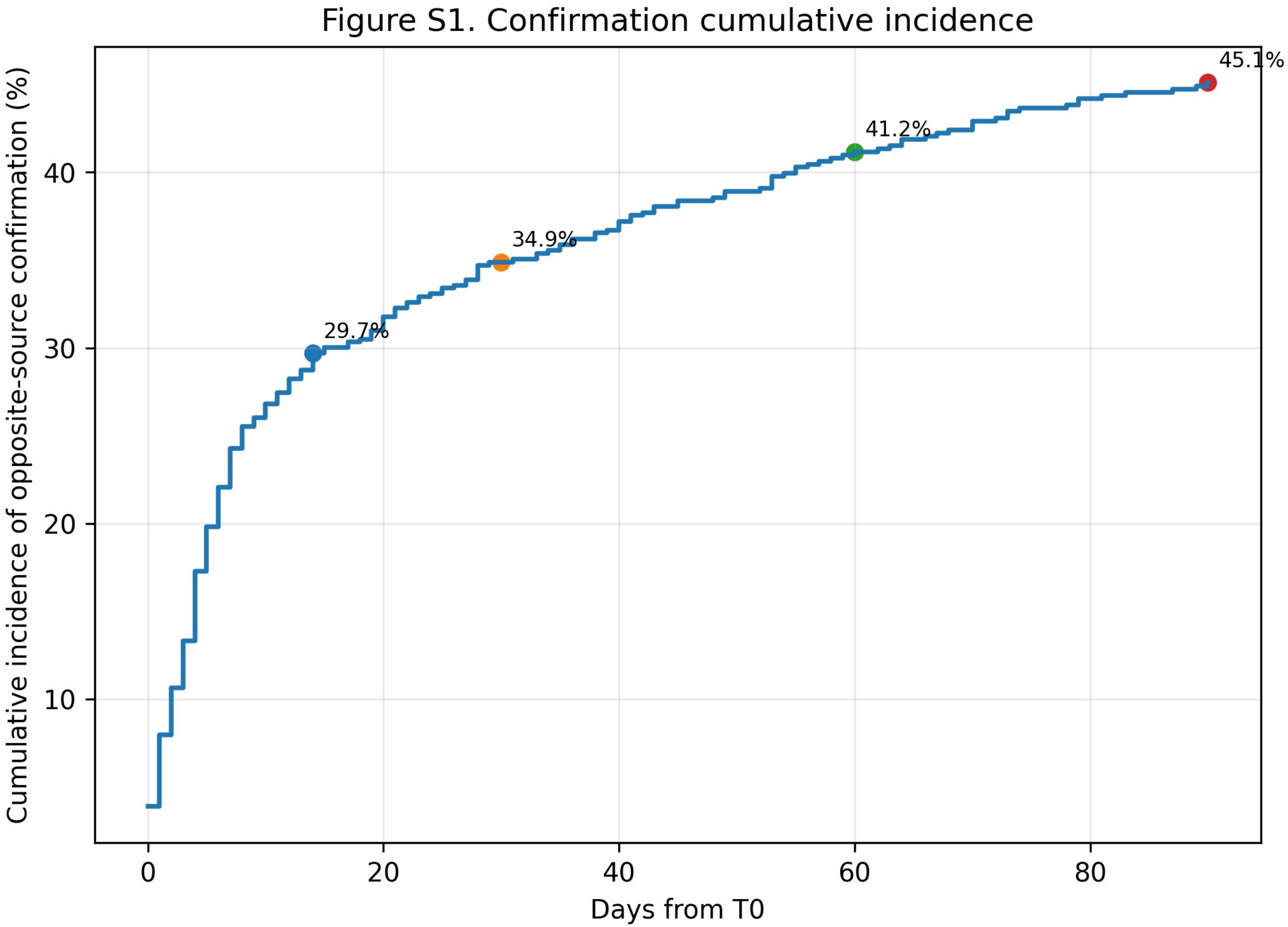


Aalen–Johansen cumulative incidence of opposite-source progression confirmation, with subsequent treatment/death as competing events and administrative close as censoring.

**Figure S2. Endpoint sensitivity battery.**

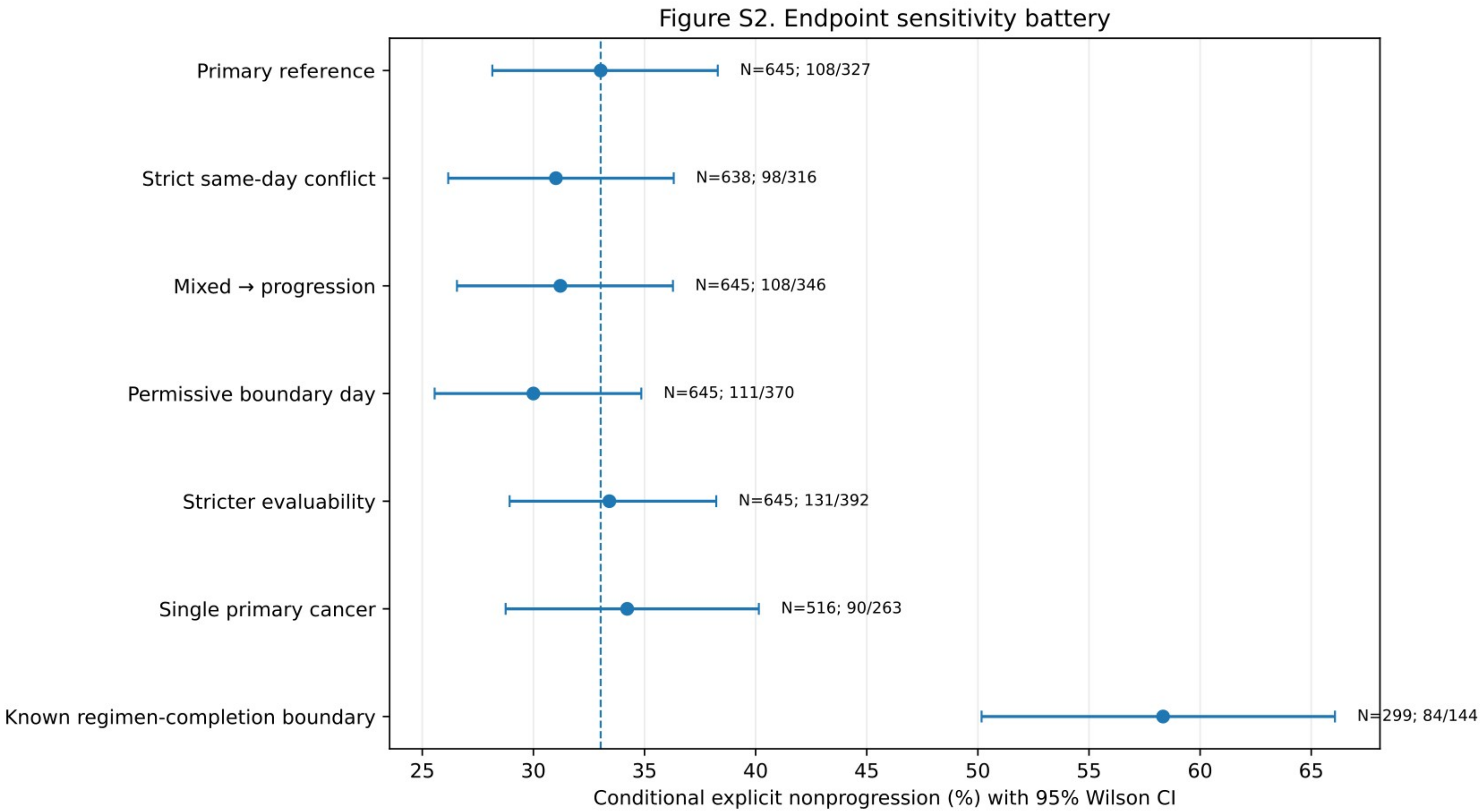


Conditional explicit nonprogression across the prespecified sensitivity set. The known-regimen-completion sensitivity (N=299; 84/144 = 58.33%) is shown without being used to redefine the primary estimand.

**Figure S3. Descriptive heterogeneity.**

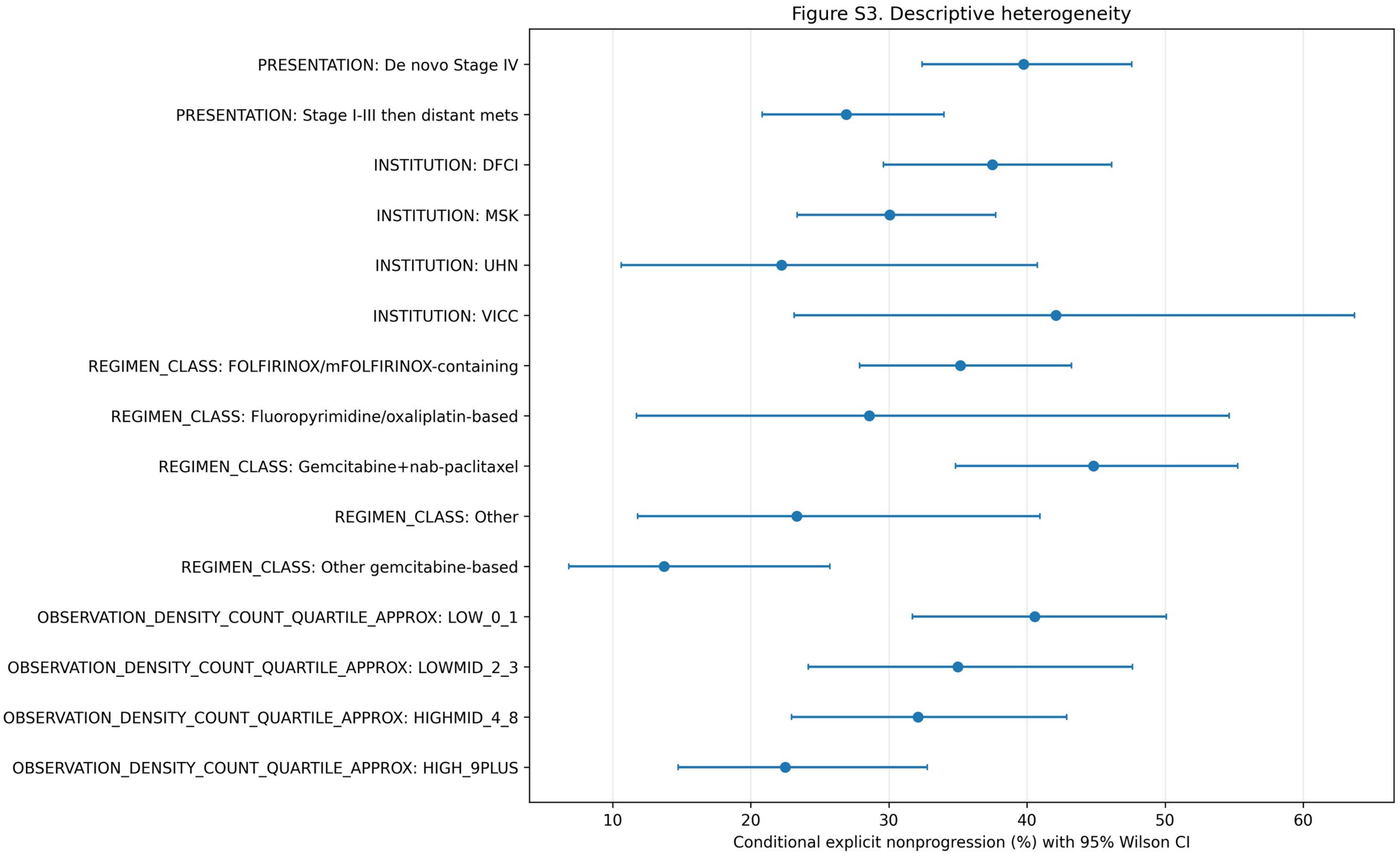


Conditional explicit nonprogression across presentation, institution, regimen class, and observation-density strata. These are descriptive/supportive analyses, not effect-modifier claims.

**Figure S4. CA19-9 window and boundary sensitivity.**

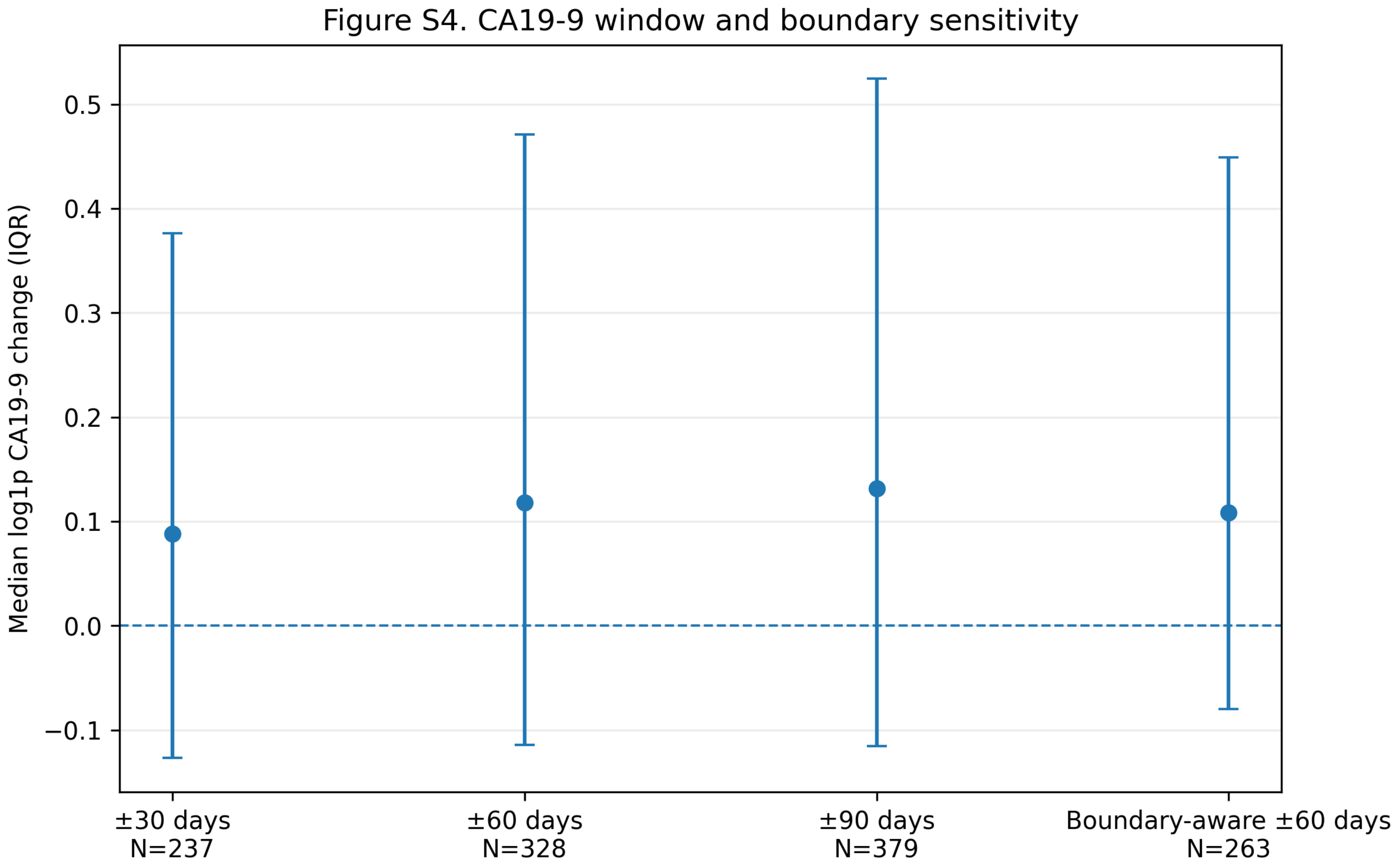


Median log1p CA19-9 change across ±30/±60/±90-day windows and the boundary-aware ±60-day sensitivity. CA19-9 remains contextual.

**Figure S5. Next-treatment cumulative incidence.**

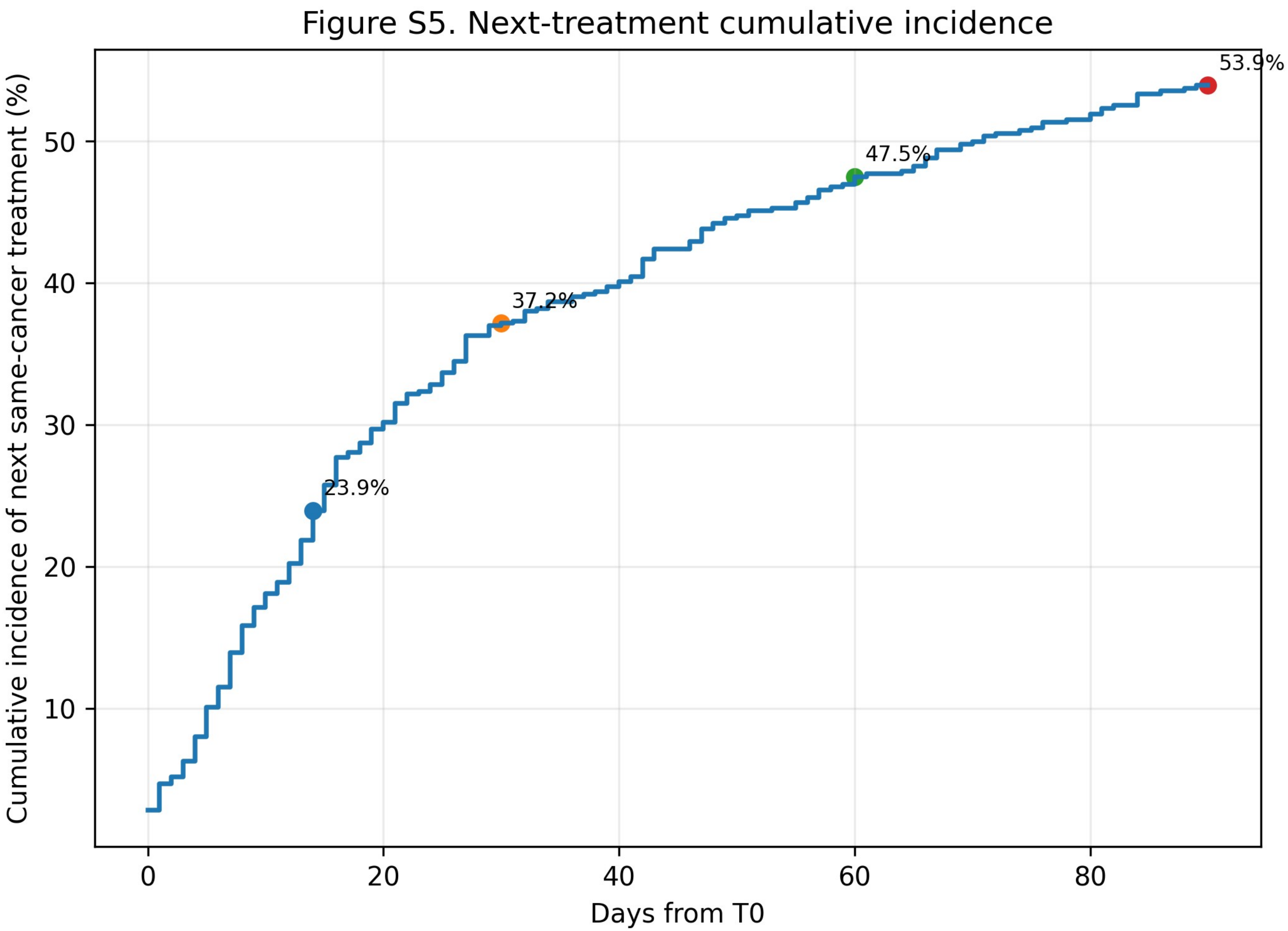


Follow-up-aware Aalen–Johansen cumulative incidence of recorded next same-cancer treatment after T0. This display is descriptive and structurally linked to the endpoint boundary.
